# The Pivotal Role of Immune Functional Assays in Deciphering Immune Function Alterations

**DOI:** 10.1101/2024.11.19.24317497

**Authors:** Marion Debombourg, Guy Oriol, Caroline Dupre, Chloé Albert-Vega, Fabienne Venet, Thomas Rimmelé, REALISM study group, Anne Conrad, Florence Ader, Vincent Alcazer, VaccHemInf study group, Karen Brengel-Pesce, Aurore Fleurie, Sophie Trouillet-Assant, William Mouton

## Abstract

Recently, immune function assessment has gained prominence in clinical settings. Immune functional assays (IFAs), involving in vitro stimulation, offer a relevant approach to complement traditional immunomonitoring methods which, while widely used, do not fully capture functional immune capabilities. Despite growing interest in IFAs, their added value remains unclear.

To address this gap, our study aimed to determine if insights from IFAs could be replicated with unstimulated immunoprofiling. Using the same analytical pipeline, we compared transcriptomic profiles (Nanostring®) between stimulated (TruCulture®) and unstimulated (PaxGene™) samples from i) patients with an overstimulated immune system 3-4 days post-sepsis onset, and ii) patients undergoing immune reconstitution 6-months post-allogeneic hematopoietic stem cell transplantation (allo-HSCT).

In sepsis, post-stimulation transcriptomic profiles revealed immune clusters linked to disease severity and outcomes, surpassing traditional markers like mHLA-DR, while baseline analyses failed to generate clinically relevant stratification. Similarly, allo-HSCT patients’ post-stimulation data revealed immune heterogeneity and treatment-related alterations not detected using baseline transcriptomic or cellular profiles alone.

Our findings emphasize the value of IFAs in uncovering functional immune alterations that unstimulated assessments may miss, which could offer deeper insights into immune dysfunction. This study supports IFAs as complementary tools to current clinical practices, enhancing patient management with a functional view of immune system dynamics.

## 2 Introduction

Over the past decade, the assessment of immune function has become a relevant approach for the evaluation of host immune capacities (1–3) and immune reconstitution quality (4,5), as well as disease monitoring (1,6,7) across various research domains and clinical contexts. In this regard, Immune Functional Assays (IFA) have emerged as essential tools (1,8), composed by an *in vitro* stimulation step followed by an analysis of the immune response induced by the stimulation, using a varying degree of analytical complexity. These analyses can range from single cytokine secretion measurement, such as interferon-gamma (9), to more intricate phenotyping techniques, such as multiplex OMIC approaches (10). The employment of IFA, and more particularly TruCulture^®^ tubes (Myriad RBM, Austin, USA) combined with Nanostring^®^ technology to quantify mRNA gene expression level, allowed us to obtain valuable insights into two distinct immunocompromised populations. In the setting of sepsis, transcriptomic data post-staphylococcal enterotoxin B (SEB) stimulation in TruCulture^®^ tubes enabled us to stratify a sepsis population according to severity and proved to be more effective than measurements of mHLA-DR, a marker classically used for immune monitoring (11). In the case of allogenic hematopoietic stem cell transplantation (allo-HSCT), gene expression observed post-SEB and post-lipopolysaccharide (LPS) stimulation allowed to identify immune functional alterations associated with ongoing immunosuppressive treatment at 6 months post-transplantation, which could not be revealed solely through cell count analyses, even though they are considered as reference markers for immune reconstitution (4).

Other studies have also highlighted the advantages of employing IFA over traditionally used immunomonitoring tools, such as peripheral white blood cell count or immune cell phenotyping conducted without any *in vitro* stimulation step (12–14). But to date, and to our knowledge, no study has addressed the question of whether a stimulation step is necessary for obtaining information that could enhance the detection of functional immune alterations. Furthermore, direct comparisons between stimulated and unstimulated samples using analytical pipelines differing only on the *in vitro* stimulation step have not been performed, thereby hindering a clear demonstration of the added value of IFA.

In this study, we aimed to address this gap by conducting two independent assessments of the added value of *in* vitro stimulation compared to baseline transcriptomic profiles for revealing immune function alterations, in the previously mentioned clinical settings: sepsis progression and immune reconstitution following allo-HSCT.

## 3 Material & Methods

### 3.1 Study population

#### Sepsis cohort

Patients with sepsis were included in the REALISM study (NCT02638779), a prospective longitudinal, single-center observational study, conducted in the anesthesiology and intensive care department of the Edouard Herriot hospital between December 2015 and June 2018 (*Hospices Civils de Lyon*, HCL, Lyon, France). Blood sampling was performed 3 to 4 days after septic shock onset. The study protocol was approved by the regional ethics committee (*Comité de Protection des Personnes Sud-Est II*, number 2015–42-2).

#### Allo-HSCT cohort

Allo-HSCT recipients were included in the prospective, single-center cohort study “VaccHemInf” (NCT03659773) between May 2018 and August 2020 at a median time of 6 months post-transplantation at the hematology department of the Lyon university hospital (HCL,France). The study protocol was approved by the regional ethics committee (*Comité de Protection des Personnes Sud-Est V*, Grenoble, France, number 69HCL17_0769).

For each study, blood samples from 10 healthy volunteers (HVs) were concomitantly obtained from the national blood service (*Etablissement Français du Sang*), following the regulatory authorizations for the handling and conservation of these samples from the regional ethics committee (*Comité de Protection des Personnes Sud-Est II*) and the French ministry of research (*Ministère de l*□*Enseignement supérieur, de la Recherche et de l*□*Innovation*, DC-2008–64).

Written informed consent was obtained from each healthy donor and from the patients or their relatives upon inclusion in these studies.

### 3.2 Sample collection

For each study, at the time of sampling, heparinized-whole blood was collected and incubated in TruCulture^®^ tubes (Myriad Rbm, Austin, TX, USA) pre-filled with either LPS or SEB, as previously described (4,11). Unstimulated whole blood samples were also collected in PaxGene^™^ Blood RNA tubes (PreAnalytiX/QIAGEN Inc., Valencia CA, USA) and stored at -80°C.

### 3.3 Isolation of RNA from PaxGene whole-blood

Total RNA was extracted from whole-blood samples using the PaxGene™ Blood RNA Kit (PreAnalytiX/QIAGEN Inc., Valencia CA, USA). After thawing, blood RNA tubes were centrifuged at 3,000g for 10 min at room temperature (RT). The supernatant was decanted, followed by the addition of 4 ml RNase-free water and redissolving of the pellet. Next, the tube was centrifuged again at 3,000 g for 10 min at RT, and the supernatant was discarded. The pellet was resuspended in 350 µl of buffer BR1 and transferred to a microcentrifuge tubeNext, 300 µl of buffer BR2 and 40 µl of proteinase K were added, mixed, and incubated for 10 min at 55°C using a shaker-incubator at 450 rpm. This lysate was transferred to a PaxGene shredder (Qiagen, Valencia, CA) spin column and centrifuged at 20,000 g for 3 min at RT. The supernatant of the flow-through fraction was transferred to a fresh microcentrifuge tube, and 350 µl of 100% ethanol was added and mixed. Then 700 µl of this sample was transferred to a PaxGene RNA spin column and centrifuged at 10,000g for 1 min at RT. The flow-through was discarded, and this step was repeated with the remaining sample from the previous step.

Next, 350 µl of buffer BR3 was added onto the RNA spin column, which was centrifuged at 10,000g for 1 min at RT. The flow-through was discarded, and 10 µL of DNase 1 was mixed with 70 µl of buffer RDD in a separate tube. This was pipetted onto the spin column membrane and incubated at 20–30°C for 15 min. Then 350 µl of buffer BR3 was added onto the RNA spin column, which was centrifuged at 10,000 g for 1 min at RT. The flow-through was then discarded. Next, 500 µl of buffer BR4 was added onto the RNA spin column, which was centrifuged at 10,000 g for 1 min at RT. The flow-through was discarded, and 500 µl of buffer BR4 was added again onto the RNA spin column and centrifuged at 10,000 g for 3 min at RT. The flow-through was discarded. The RNA spin column was transferred to a 1.5-ml microcentrifuge tube, and 40 µl of buffer BR5 was pipetted onto the column to elute the membrane-bound RNA. The column was then centrifuged at 10,000 g for 1 min at RT. This step was repeated with the same 40-µl buffer BR5, and the eluate was incubated at 65°C for 5 min, then chilled on ice. RNA concentration was estimated using the Nanodrop One spectrophotometer (Thermo Scientific, Swedesboro, NJ) according to the manufacturer’s instruction.

### 3.4 Gene expression analysis

Gene expression was evaluated through a 89-(11) and 144-gene panel (4) using the NanoString^®^ technology for the sepsis and allo-HSCT study, respectively. Briefly, 300 ng of RNA were hybridized to the probes at 67 °C for 18 h using a thermocycler (Biometra, Tprofesssional TRIO, Analytik Jena AG, Jena, Germany). After removal of excessive probes, samples were loaded into the nCounter Prep Station (NanoString^®^ Technologies, Seattle, WA, USA) for purification and immobilization onto the internal surface of a sample cartridge for 2–3 h. The sample cartridge was then transferred and imaged on the nCounter Digital Analyzer (NanoString^®^ Technologies) where color codes were counted and tabulated for each panel of genes. Data treatment and normalization were performed on nSolver^®^ analysis software (version 4.0, NanoString^®^ Technology) using internal controls and 3 housekeeping genes (detailed in **Table S1 and S2**). Of note, for both cohorts, analyses were conducted using genes with expressions exceeding the background noise threshold in more than 75% of individuals, as previously described (4,11).

### 3.5 Statistical Analysis

Normality testing was performed using the Shapiro-Wilk normality test. The distribution of quantitative data was expressed as mean (range) or median (interquartile range, [IQR]) where appropriate. Cluster analyses were performed using the PAM method with correlation and average distance, as previously described by Albert-Vega et *al*. (11). The alluvial plot was obtained via http://www.bioinformatics.com.cn, a free online platform for data analysis and visualization. Principal component analysis (PCA) was carried out using Partek Genomics Suite software (version 7.0; Partek Inc., St. Louis, MO), and Euclidean distances were calculated as previously described by Mouton et *al*. (4). Differences in Euclidean distances between groups were calculated using a non-parametric unpaired Wilcoxon test with Benjamini correction. Differences in standard deviations (SD) were calculated using paired Pitman-Morgan test. Statistical analyses were conducted using GraphPad Prism software (version 8; GraphPad software, La Jolla, CA, USA) and R (version R.2.2). *P* values and adjusted *P* values (*P* adj) < 0.05 were considered significant.

## 4 Results

### 4.1 Immune function assessment appears as an essential tool to obtain clinically relevant clustering in the context of sepsis

Overall, 28 out of the 30 patients with sepsis as well as 10 HVs initially included in the Albert-Vega et *al*. study were analyzed herein, as 2 unstimulated samples had not been collected. Among these patients, 7 developed a hospital-acquired infection (HAI) during ICU stay, and 3 had died at day 28 **(Table S3)**. Following the data control and normalization steps described above, 81 out of 86 genes initially analyzed were kept for analyses through PaxGene™ tubes. To evaluate the added value of the stimulation in identifying immune functional alterations in a sepsis population, statistical analyses and clustering must be conducted using an equal number of patients and genes, whose expression was measured post-stimulation or from PaxGene™ tubes without stimulation. To that end, we first aimed to validate that the conclusions derived from the restricted post-stimulation dataset were consistent with those obtained from the complete dataset published in the Albert-Vega et *al*. study.

We thus conducted again the multivariate clustering analysis from gene expression levels post-SEB stimulation using 81 genes, 28 patients with sepsis, and 10 HVs. The analysis found 3 clusters with the same composition as previously described by Albert-Vega et *al*. (**Figure 1 left, Table S4**). The first cluster (n=11) grouped together all the HVs and one patient with sepsis, constituting the healthier cluster, gathering immunocompetent individuals. The second cluster (n=13) included all non-survivors, hence designated as the severe cluster. It was characterized by a diminished immune responsiveness upon *in vitro* SEB stimulation and a specific modulation of genes previously described to be associated with mortality, such as MDC1 and IFIT44L. The third cluster (n=14) included 86% (6/7) of the patients with sepsis who developed a HAI, forming the intermediate cluster. As demonstrated by Albert-Vega et *al*., these patients exhibited, among other, an upregulation of the HLA family and interferon-related genes, suggesting a potential for immune recovery, implying that patients identified in this cluster may benefit from immunostimulatory therapy.

**FIGURE 1.**
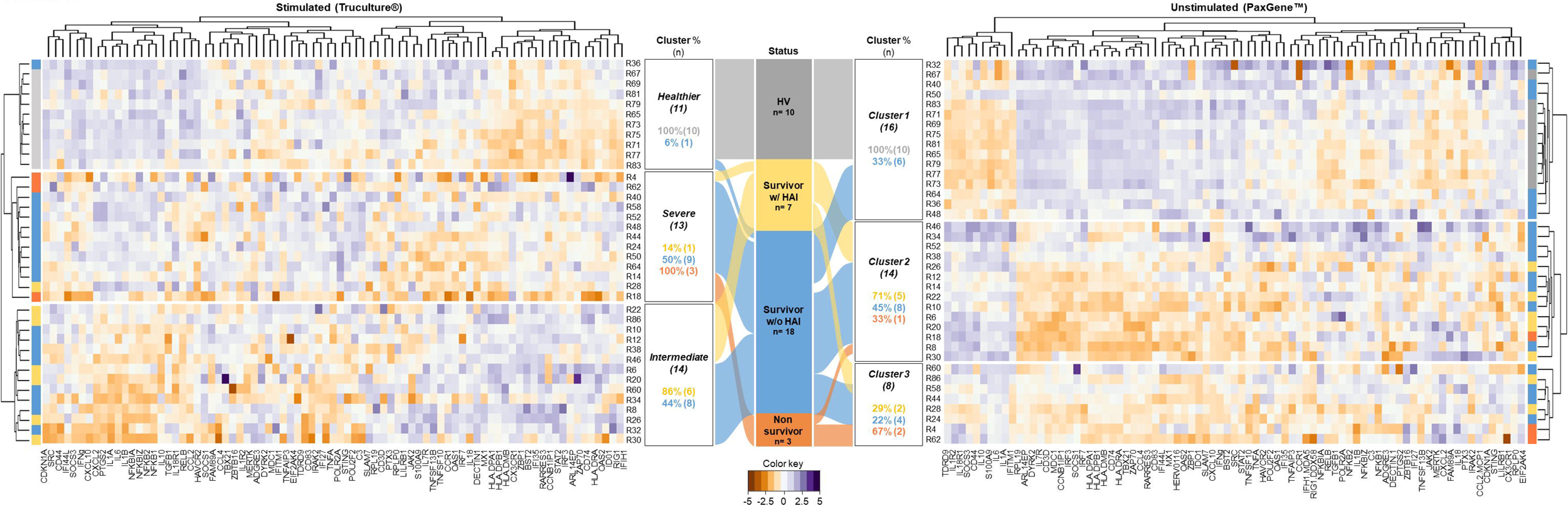

Having confirmed the consistency of the conclusions that can be drawn from the restricted dataset, we conducted the same analyses on PaxGene™ tubes (**Figure S1.A**). Using the dataset obtained from PaxGene™ tubes, we identified an equivalent of the healthier cluster comprising all HVs, consistent with observations from the post-stimulation gene dataset. However, in the absence of stimulation, this healthier cluster included 4 of 9 patients with sepsis who had been classified within the severe cluster identified post-SEB stimulation, including patients who exhibited an evident and profound alteration of immune function post-stimulation **(Figure 1 right)**. This discrepancy demonstrates the loss of ability to distinguish immunocompetent individuals among a sepsis population based on gene expression analysis from unstimulated whole blood.

Altogether, these results highlight a notable discrepancy in patient stratification according to immune alteration profiles when determined with or without *in vitro* stimulation, and suggest that, during the course of sepsis, immune function assessment can reveal distinct immune profiles that are coherent with clinical characteristics.

### 4.2 Immune function assessment remains necessary in the context of allo-HSCT to uncover immune function alterations

We then evaluated the added value of the stimulation to reveal immune functional alterations in another clinical context, e.g. during the immune reconstitution after allo-HSCT. For this study, 59 of the 60 allo-HSCT recipients and 5 of the 10 HVs initially included were analyzed, as one patient sample did not pass quality control, and no PaxGene™ tubes were available for 5 HVs. Regarding the hematological- and transplant-related characteristics of allo-HSCT recipients included at a median [IQR] of 6.5 [5.8-8.3] months after transplantation, 52.5% had been transplanted due to acute myeloid leukemia, 35.6% had active graft-versus-host disease (GvHD) at inclusion, and 32.2% were undergoing immunosuppressive treatment at inclusion **(Table S5)**.

Following the data control and normalization steps, 121 out of 138 and 134 genes initially analyzed post-SEB and -LPS stimulation, respectively, were kept for analyses through PaxGene™ tubes. As for the sepsis study, we first aimed to confirm whether the same conclusions obtained from the entire post-stimulation transcriptomic dataset used in the Mouton et *al*. study could be replicated using the new restricted dataset.

We thus conducted again the PCA from gene expression levels post-stimulation with either LPS or SEB using 121 genes, 59 allo-HSCT recipients, and 5 HVs. Similarly, post-stimulation gene expression analysis revealed a strong homogeneity among HVs, while allo-HSCT recipients represented a more heterogeneous population (**Figure S2**). For quantitative purposes, we once again calculated, using the PCA projection from the restricted dataset, the Euclidean distance of each allo-HSCT recipient to the centroid of the HV population, which serves as a reference value for a functional immune response. We hypothesized that a greater distance corresponds to a more impaired immune response and assessed the association between these distances and clinical data. Thus, using this restricted post-stimulation dataset, we were able to confirm an increase in Euclidean distance associated with an ongoing immunosuppressive treatment (median [IQR] 9.71 [4.91-12.6] versus 13.77 [11.34-18.43] *P* adj < 0.047, **Table S6**) (4).

We then proceeded to conduct the same analyses on PaxGene™ tubes **(Figure S1.B)**. Whether obtained post-stimulation or under unstimulated conditions, the immune profiles projected onto the first 2 principal components of the PCA revealed similar overall variability, explaining 36.6% and 36.7% of the overall variance, respectively (**Figure 2.A**). Nevertheless, as illustrated in the PCA overlay, the immune functional heterogeneity of allo-HSCT recipients, captured using gene expression analysis through unstimulated whole blood, was less pronounced. This was supported by two indicators, the inertia of the point cloud formed by the allo-HSCT immune profiles and the SD of the Euclidean distance **(Figure 2.B)**, which were both highly reduced in the unstimulated condition compared to the stimulated one (inertia: 39.4 *vs*. 87.5 and SD: 3.814 *vs*. 6.977, p-value <0.001). In addition, neither ongoing immunosuppressive treatment (**Figure 2.C**) nor any other parameter (**Table S7**) could account for the significant increase in the Euclidean distance using the gene expression dataset obtained without whole blood stimulation, in contrast to that obtained post stimulation. It appears that in the context of allo-HSCT, immune function assessment through IFA uncovers a treatment-dependent immune response, which could help improve the monitoring of post-transplantation immune reconstitution by complementing the classical markers currently used.

**FIGURE 2.**
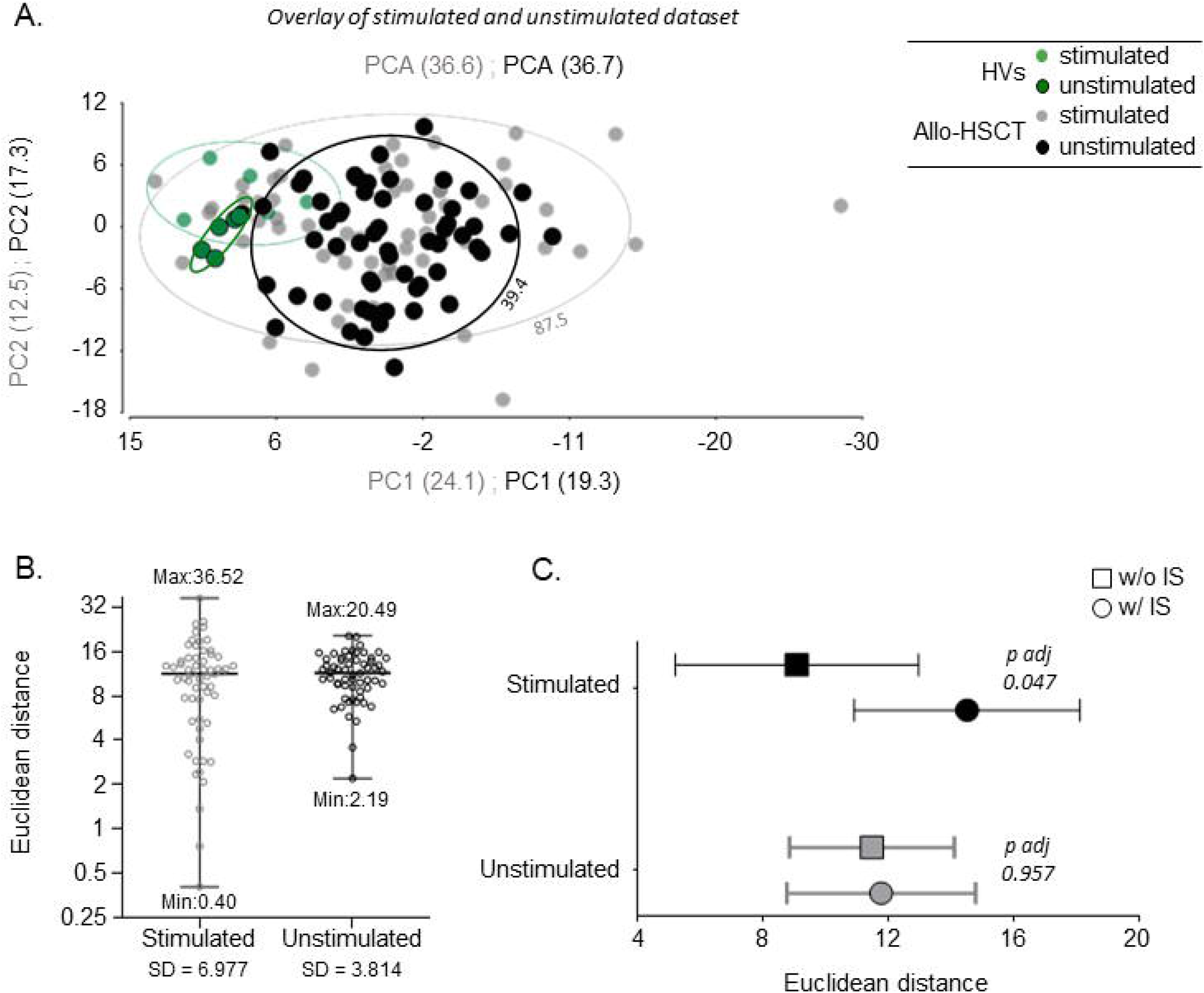

## 5 Discussion

In this study, we aim to evaluate the added value of IFA in capturing altered immune function compared to unstimulated assays in two distinct clinical contexts. To do so, we compared transcriptomic data obtained after a non-specific whole-blood stimulation in TruCulture® with those acquired using PaxGene samples, employing the same analytical pipeline.

In the first context of sepsis immune monitoring, several basal-state biomarkers, i.e., without stimulation step, are routinely used in clinical practice, such as mHLA-DR, CD4^+^/CD8^+^ ratio, and circulating IL-10 (15). Indeed, persistent low mHLA-DR expression (16–18), decrease in CD4^+^/CD8^+^ ratio (19–22) as well as an increase circulation in IL-10 (23) have been reported to predict mortality in septic shock. Several studies have already employed IFA to assess functional immunity in this context; as Antonakos et *al*. who demonstrated that TNF-α production post-LPS stimulation on day 3 post-sepsis onset could discriminate patients with sepsis from healthy control subjects (24), or Mazer et *al*. who used IFN-γ and TNF-α ELISpot assays post-anti-CD3/anti-CD28 antibodies and LPS stimulation, respectively, to depict early, profound and sustained suppression of functional immunity in deceased patients (25). Despite the interest in IFA for assessing immune function, no study has yet clearly assessed the added value of stimulation compared to basal-state biomarkers in depicting immune alterations. Therefore, using the same analytical pipeline employed by Albert-Vega et *al*. (11), which demonstrated relevant stratification based on immune functional profiles during the course of sepsis, we evaluated whether the dataset obtained from unstimulated PaxGene samples collected from the same patients at the same visit could reveal similar results. We observed that the unstimulated dataset did not yield clinically relevant clusters, underscoring the value of stimulation in revealing distinct immune profiles during sepsis.

Moreover, our results align with the previous observation made by Albert-Vega et *al*., showing that patients’ stratification according to post-SEB stimulation transcriptomic profiles was more effective than using the commonly employed mHLA-DR marker to underline the heterogeneity in sepsis. This reinforces the relevance of IFA employment in this setting, as also suggested by Wang et *al*. (6).

In the second context of allo-HSCT, immune cell counts such as TCD4^+^ cell count and CD4^+^/CD8^+^ ratio are classically quantified to monitor immune reconstitution post-transplant (26,27). However, despite their widespread clinical use, these approaches provide no information regarding the qualitative characteristics of this immune reconstitution (14,28). In this regard, IFA have demonstrated complementary value to these classically used methods. Gjaerde et *al*. conducted proteomic analysis following whole-blood stimulation in TruCulture®, revealing heterogeneity in cytokine production among patients and identifying a cluster with reduced responses, suggesting possible functional immune deficiency (5). Similarly, Mouton et *al*. used non-specific TruCulture® stimulation coupled with a transcriptomic approach to capture a broad range of immune profiles, identifying altered immune functional profiles associated with ongoing immunosuppressive treatment (4). As for the sepsis context, we aimed to distinctly evaluate the added-value of IFA compared to basal-state biomarkers in deciphering immune alterations. To do so, using the same analytical pipeline as Mouton et *al*., we analyzed unstimulated PaxGene sample collected from the same patients at the same visit. In comparison with the results obtained post *in vitro* stimulation, the use of unstimulated dataset failed to detect any functional alterations, resulting in homogeneous transcriptomic immune profiles among patients. These profiles were not associated with any clinical event or characteristic, especially with the use of immunosuppressive treatments, contrasting with the findings observed post-stimulation. Once again, these results were in line with conclusion made by Mouton et *al*., which underlined that a clustering approach post-stimulation using transcriptomic data is more effective than solely analyzing cell counts in revealing the heterogeneity of immune profiles during post-transplant reconstitution. This supports the relevance of IFA, as also suggested by Naik et *al*. (14).

However, this study has limitations that need to be addressed. Studies with larger sample sizes will be essential to fully evaluate the clinical utility of IFAs and their potential benefits for patient care. Here, we used previously established bioinformatics pipelines specifically designed for post-stimulation data to enable a precise comparison; however, we acknowledge that alternative methods may be more suitable for unstimulated datasets. Finally, studies incorporating longitudinal follow-ups of immunocompromised patients at various stages of their conditions would be of interest.

Overall, the present analyses showed that the conclusions obtained through a clustering-based stratification of post-stimulation data, in two different clinical contexts could not be replicated using unstimulated samples. We reinforce the interest of IFA as complementary tool to traditional immunomonitoring methods, as already well demonstrated for specific immunity in infectious contexts, such as SARS-CoV-2 (29) or *Mycobacterium tuberculosis* (30). The design of the present study highlighted, for the first time, the added value of the stimulation step in identifying functional immune alterations. This observation could pave the way for, or at least encourage, the broader implementation of IFA as a complementary tool in immunomonitoring.

## Data Availability

all data in the present study are available upon reasonable request to the authors

## 7 Author Contributions

All authors were involved in the analysis and interpretation of data, as well as drafting the manuscript or revising it critically for important intellectual content. MD, STA and WM made substantial contributions to the conception and design of the study and designed the experiments. MD and WM performed the experiments. MD, GO, CD, and WM performed the data analyses. MD, WM and STA drafted the manuscript. All authors read and approved the final manuscript. WM and STA take responsibility for the integrity of the data analysis.

## 8 Conflict of Interest

MD, GO, KBP and AF are employed by the in-vitro diagnostic company bioMérieux.

## 9 Funding

*REALISM study* - This work was supported by the French National Research Agency through a grant awarded to BIOASTER (Grant number #ANR-10-AIRT-03) and from bioMérieux, Sanofi and GSK.

*VaccHemInf – FIGHT study* – This work was supported by an internal grant from the *Hospices Civils de Lyon* (Appel d’Offre Jeune Chercheur 2018, to A.C.); the *Région Auvergne-Rhône-Alpes* (Pack Ambition Recherche 2019, to F.A.)

This work was supported by the public grant overseen by the French National Research Agency (C*haires industrielles 2023)* and supported by the *Association Nationale de la Recherche et de la Technologie* (ANRT) with a CIFRE fellowship granted to Marion Debombourg.

## 10 Acknowledgments

The authors thank Jonathan Lopez, Pauline Berlier and Isabelle Mosnier for their technical assistance on Nanostring molecular biology; Verena Landel for language editing and critical reading of the manuscript.

## Notes

### Competing Interest Statement

The authors have declared no competing interest.

### Author Declarations

The REALISM study protocol was approved by the regional ethics committee (Comite de Protection des Personnes Sud-Est II, number 2015 42 2). The VaccHemInf protocol was approved by the regional ethics committee (Comite de Protection des Personnes Sud-Est V, Grenoble, France, number 69HCL17 0769).

