## Supplementary data for "The Pivotal Role of Immune Functional Assays in Deciphering Immune Function Alterations"

Supplementary Material

### 1 Supplementary Tables

**Supplementary Table S1. Nanostring targeted genes and accession numbers for the sepsis study, based on the Albert-Vega et al. study** (1)**.**

| Target genes | Accession number |
| --- | --- |
| *ADGRE3* | NM_032571.2 |
| *ARL14EP* | NM_152316.1 |
| *BST2* | NM_004335.2 |
| *C3* | NM_000064.2 |
| *CCL2* | NM_002982.3 |
| *CCL4* | NM_002984.2 |
| *CCNB1IP1* | NM_182849.2 |
| *CCR1* | NM_001295.2 |
| *CD3D* | NM_000732.4 |
| *CD44* | NM_001001392.1 |
| *CD74* | NM_001025159.1 |
| *CD83* | NM_004233.3 |
| *CDKN1A* | NM_000389.2 |
| *CLEC7A/DECTIN1* | NM_197954.2 |
| *CX3CR1* | NM_001337.3 |
| *CXCL10/IP10* | NM_001565.1 |
| *CXCL2/MIP2A* | NM_002089.3 |
| ***DECR1*** | **NM_001359.1** |
| *DDX58/RIG1* | NM_014314.3 |
| *DYRK2* | NM_003583.3 |
| *EIF2AK4* | NM_001013703.2 |
| *FAM89A* | NM_198552.2 |
| *HAVCR2/TIM3* | NM_032782.3 |
| *HLA-DMB* | NM_002118.3 |
| *HLA-DPA1* | NM_033554.2 |
| *HLA-DPB1* | NM_002121.4 |
| *HLA-DRA* | NM_019111.3 |
| ***HPRT1*** | **NM_000194.1** |
| *IDO1* | NM_002164.3 |
| *IFI27* | NM_005532.3 |
| *IFI35* | NM_005533.3 |
| *IFI44L* | NM_006820.2 |
| *IFIH1* | NM_022168.2 |
| *IFITM1* | NM_003641.3 |
| *IFNG* | NM_000619.2 |
| *IL10* | NM_000572.2 |
| *IL18* | NM_001562.2 |
| *IL18R1* | NM_003855.2 |
| *IL1A* | NM_000575.3 |
| *IL1B* | NM_000576.2 |
| *IL1R2* | NM_004633.3 |
| *IL6* | NM_000600.1 |
| *IL7R* | NM_002185.2 |
| *IRAK2* | NM_001570.3 |
| *IRF3* | NM_001571.5 |
| *IRF7* | NM_001572.3 |
| *JAK2* | NM_004972.2 |
| *LILRB1* | NM_001081637.1 |
| *MDC1* | NM_014641.2 |
| *MERTK* | NM_006343.2 |
| *MX1* | NM_002462.2 |
| *NFKB1* | NM_003998.2 |
| *NFKB2* | NM_002502.2 |
| *NFKBIA* | NM_020529.1 |
| *NFKBIZ* | NM_001005474.1 |
| *OAS1* | NM_001032409.1 |
| *OAS2* | NM_016817.2 |
| *POLR2A* | NM_000937.2 |
| *POU2F2* | NM_002698.2 |
| *PTGS2* | NM_000963.1 |
| *PTX3* | NM_002852.3 |
| *RARRES3* | NM_004585.3 |
| *RELB* | NM_006509.2 |
| *RPL19* | NM_000981.3 |
| *RPLP0* | NM_001002.3 |
| *S100A9* | NM_002965.2 |
| *SLAMF7* | NM_021181.3 |
| *SOCS1* | NM_003745.1 |
| *SOCS3* | NM_003955.3 |
| *SRC* | NM_005417.3 |
| *STAT2* | NM_005419.2 |
| *TBX21* | NM_013351.1 |
| ***TBP*** | **NM_001172085.1** |
| *TDRD9* | NM_153046.2 |
| *TGFB1* | NM_000660.3 |
| *TMEM173/STING* | NM_198282.1 |
| *TNFA* | NM_000594.2 |
| *TNFAIP3* | NM_006290.2 |
| *TNFSF10* | NM_003810.2 |
| *TNFSF13B* | NM_006573.4 |
| *ZAP70* | NM_001079.3 |
| *ZBP1* | NM_001160419.2 |
| *ZBTB16* | NM_006006.4 |
| *121601901-HERV0116* | chr12:112972627-112975754 |

Housekeeping genes used for the gene expression analysis are depicted in bold

**Supplementary Table S2. Nanostring targeted genes and accession numbers for the allo-HSCT study, based on the Mouton et al. study** (2)**.**

| **Gene Name** | **Accession number** |
| --- | --- |
| *ADGRE3* | NM_032571.2 |
| *B2M* | NM_004048.2 |
| *BANK1* | NM_001083907 |
| *BATF* | NM_006399.3 |
| *BATF3* | NM_018664.2 |
| *BCL2* | NM_000657.2 |
| *BST2* | NM_004335.2 |
| *C1QB* | NM_000491.3 |
| *C3* | NM_000064.2 |
| *CCL23* | NM_145898.1 |
| *CCL3* | NM_002983.2 |
| *CCL4* | NM_002984.2 |
| *CCR1/ RANTES R* | NM_001295.2 |
| *CCR5* | NM_000579.1 |
| *CCR7* | NM_001838.2 |
| *CCRL2* | NM_003965.4 |
| *CD209/DC-SIGN* | NM_021155.2 |
| *CD244* | NM_016382.2 |
| *CD27 / TNFRSF7* | NM_001242.4 |
| *CD274* | NM_014143.3 |
| *CD4* | NM_000616.4 |
| *CD40* | NM_001250.4 |
| *CD44* | NM_001001392.1 |
| *CD45RA* | NM_002838.4 |
| *CD79A* | NM_001783.3 |
| *CD79B* | NM_021602.2 |
| *CD80* | NM_005191.3 |
| *CD83* | NM_004233.3 |
| *CDKN1A* | NM_000389.2 |
| *CEBPB* | NM_005194.2 |
| *CSF1* | NM_000757.4 |
| *CSF2* | NM_000758.2 |
| *CSF2RB* | NM_000395.2 |
| *CX3CR1* | NM_001337.3 |
| *CXCL10/IP10* | NM_001565.1 |
| *CXCL11* | NM_005409.4 |
| *CXCL2/MIP2alpha* | NM_002089.3 |
| *CXCL9* | NM_002416.1 |
| *CXCR1* | NM_000634.2 |
| ***DECR1*** | **NM_001359.1** |
| *FCGR2B* | NM_001002273.1 |
| *FCRL5* | NM_031281 |
| *FCRLA* | NM_032738 |
| *FOXP3* | NM_014009.3 |
| *GBP1* | NM_002053.1 |
| *GBP5* | NM_052942.3 |
| *GZMB* | NM_004131.3 |
| *HAVCR2* | NM_032782.3 |
| *HLA-DMB* | NM_002118.3 |
| *HLA-DPA1* | NM_033554.2 |
| *HLA-DPB1* | NM_002121.4 |
| *HLA-DRA* | NM_019111.3 |
| ***HPRT1*** | **NM_000194.1** |
| *ICAM1* | NM_000201.2 |
| *IDO1* | NM_002164.3 |
| *IFIH1/MDA5* | NM_022168.2 |
| *IFIT2* | NM_001547.4 |
| *IFITM1* | NM_003641.3 |
| *IFNG* | NM_000619.2 |
| *IGHG1* | NM_001040077 |
| *IL1B* | NM_000576.2 |
| *IL1RN* | NM_000577.3 |
| *IL27* | NM_145659.3 |
| *IL2RA* | NM_000417.1 |
| *IL8* | NM_000584.2 |
| *IL9* | NM_000590.1 |
| *IRAK2* | NM_001570.3 |
| *IRF4* | NM_002460.1 |
| *IRF5* | NM_002200.3 |
| *IRF7* | NM_001572.3 |
| *JAK2* | NM_004972.2 |
| *LAG3* | NM_002286.5 |
| *LAMP3* | NM_014398.3 |
| *LIF* | NM_002309.3 |
| *LILRA3* | NM_006865.3 |
| *LILRA5* | NM_181879.2 |
| *LILRB1* | NM_032571.2 |
| *LILRB4* | NM_012276.3 |
| *LTA* | NM_000595.2 |
| *MME* | NM_000902.2 |
| *MS4A1 / CD20* | NM_152866.2 |
| *MX1* | NM_002462.2 |
| *NFKB1* | NM_003998.2 |
| *NFKB2* | NM_002502.2 |
| *NFKBIA* | NM_020529.1 |
| *NFKBIZ* | NM_001005474.1 |
| *PDCD1LG2* | NM_025239.3 |
| *PECAM1* | NM_000442.3 |
| *PML* | NM_002675.3 |
| ***POLR2A*** | **NM_000937.2** |
| *POU2AF1* | NM_006235 |
| *POU2F2* | NM_002698.2 |
| *PPIB* | NM_000942.4 |
| *PTX3* | NM_002852.3 |
| *RARRES3* | NM_004585.3 |
| *RELB* | NM_006509.2 |
| *RPL19* | NM_000981.3 |
| *SERPING1* | NM_000062.2 |
| *SLAMF1* | NM_003037.2 |
| *SLAMF7* | NM_021181.3 |
| *SOCS1* | NM_003745.1 |
| *SOCS3* | NM_003955.3 |
| *SPP1* | NM_000582.2 |
| *SRC* | NM_005417.3 |
| *STAT1* | NM_007315.2 |
| *STAT2* | NM_005419.2 |
| *TAGAP* | NM_054114.3 |
| *TAP1* | NM_000593.5 |
| *TAP2* | NM_000544.3 |
| *TBP* | NM_001172085.1 |
| *TCF7* | NM_003202.2 |
| *TLR7* | NM_016562.3 |
| *TNFA* | NM_000594.2 |
| *TNFAIP3* | NM_006290.2 |
| *TNFAIP6* | NM_007115.2 |
| *TNFRSF17/BCMA* | NM_001192.2 |
| *TNFRSF4* | NM_003327.2 |
| *TNFRSF9* | NM_001561.4 |
| *TNFSF10* | NM_003810.2 |
| *TNFSF13B* | NM_006573.4 |
| *TNFSF15* | NM_001204344.1 |
| *TRAF1* | NM_005658.3 |
| *XCL1* | NM_002995.1 |
| *ZAP70* | NM_001079.3 |

Housekeeping genes used for the gene expression analysis are depicted in bold

**Supplementary table S3. Clinical data for patients with sepsis**

|  | **Patients with sepsis (n=28)** |
| --- | --- |
| **Admission data** |  |
| Sex, male, n (%) | 20 (71.4) |
| Median age, years [IQR] | 66.00 [60.00-78.25] |
| Median BMI, kg/m2 [IQR] | 24.61 [21.79-26.58] |
| Median SAPS II [IQR] | 47.00 [39.50-55.25] |
| Median SOFA score (day 1) [IQR] | 9.00 [7.75-10.00] |
| Mechanical ventilation, n (%) | 19 (67.9) |
| Median plasma lactate level, mM [IQR] | 2.20 [1.70-2.80] |
| Shock, n (%) | 19 (67.9) |
| Median CCI [IQR] | 2.00 [0.75-4.00] |
| *Comorbidities^a^, n (%)* |  |
| 0 | 7 (25.0) |
| ≥ 1 | 21 (75.0) |
| *Primary site of infection, n (%)* |  |
| Abdominal | 11 (39.3) |
| UTI | 2 (7.1) |
| SST | 2 (7.1) |
| Pulmonary | 10 (35.7) |
| Others | 3 (10.8) |
| *Type of primary infection, n (%)* |  |
| Community-acquired | 21 (75.0) |
| Hospital-acquired | 7 (25.0) |
| *Documentation of infection, n (%)* |  |
| Gram-negative | 5 (17.9) |
| Gram-positive | 10 (35.7) |
| Virus | 0 |
| Fungal | 4 (14.3) |
| Co-infection | 3 (10.7) |
| Non-documented infection | 6 (21.4) |
| Hydrocortisone, n (%) | 7 (25.0) |
| **Day 3–4 data** |  |
| *Immunology* |  |
| Median mHLA-DR, Ab/C [IQR] | 4511.32 [3125.37-8053.97] |
| Median TNFα secretion post-LPS stimulation, pg/mL [IQR] | 1089.78 [669.71-1733.68] |
| **Outcomes** |  |
| Vasopressor requirement, n (%) | 26 (92.9) |
| Median vasopressor duration, days [IQR] | 1.91 [1.04-3.34] |
| Hospital-acquired infection, n (%) | 7 (25.0) |
| Median ICU length of stay, days [IQR] | 9.00 [6.50-13.00] |
| Missing data | 1 |
| Median hospital length of stay, days [IQR] | 28.00 [13.25-43.50] |
| Missing data | 3 |
| Mortality at day 28, n (%) | 3 (10.7) |

SAPS II was calculated after admission and SOFA score was measured after 24 h of ICU stay.

^a^: Presence of comorbidities was affirmative when at least one of the following comorbidity was present in the patient: chronic pulmonary disease, heart failure, myocardial infarction, ulcer, diabetes, renal failure, or malignant solid tumor.

*Abbreviations*: *BMI*, body mass index; *SAPS II*, simplified acute physiology score; *SOFA*, sequential organ failure assessment; *CCI*, Charlson comorbidity index; *UTI*, urinary tract infection; *SST*, skin and soft tissue; *HLA-DR*, human leukocyte antigen DR; *TNFα*, tumor necrosis factor alpha; *LPS*, lipopolysaccharide; *ICU*, intensive care unit

**Supplementary Table S4. Comparison of the individual composition of the clusters obtained from stimulated TruCulture analysis before and after removal of patients and genes from the original published sepsis cohort** (1)**.**

|  | | **TruCulture new dataset** | | |
| --- | --- | --- | --- | --- |
| **TruCulture published data** |  | **Cluster 1** | **Cluster 2** | **Cluster 3** |
|  | **Cluster 1** |  |  | R81; R83; R65; R67; R69; R71; R73; R75; R77; R79; R36 |
|  | **Cluster 2** | R32; R10; R12; R34; R38; R46; R8; R60; R20; R22; R26; R30; R6; R86 |  |  |
|  | **Cluster 3** |  | R14; R18; R24; R28; R4; R40; R44; R48; R50; R52; R58; R62; R64 |  |

HV are depicted in blue, patients with sepsis in black, non-survivors in red, and hospital-acquired infections in orange. There was no difference in the clustering between the two datasets.

**Supplementary Table S5. Clinical data in allo-HSCT recipients**

|  | **Allo-HSCT recipients (n=59)** |
| --- | --- |
| **Demographics** |  |
| Age, median [IQR] | 44 [33.5-60.5] |
| Male, n (%) | 34 (57.6) |
| **Time from transplantation, months (median [IQR])** | 6.5 [5.8-8.3] |
| **Hematological and transplant-related characteristics, n (%)** |  |
| *Underlying hematological disease* |  |
| Acute myeloid leukemia and related neoplasms | 31 (52.5) |
| Myelodysplastic syndromes | 7 (11.9) |
| Myeloproliferative neoplasms | 1 (1.7) |
| B-lymphoblastic leukemia/lymphoma | 11 (18.6) |
| T-lymphoblastic leukemia/lymphoma | 2 (3.4) |
| Mature neoplasms: T, NK, or B cells | 3 (5.1) |
| Hodgkin lymphoma | 1 (1.7) |
| Others | 3 (5.1) |
| *CR before the engraftment* | 53 (89.8) |
| *Donor types* |  |
| Geno-identical | 18 (30.5) |
| Haplo-identical | 11 (18.6) |
| Pheno-identical | 29 (49.2) |
| Fully matched | 23 (79.3) |
| HLA mismatched | 6 (20.7) |
| *Stem cell source* |  |
| Peripheral blood cells | 47 (79.7) |
| Bone marrow | 11 (18.6) |
| Cord blood | 1 (1.7) |
| *Conditioning regimen* |  |
| MAC | 37 (62.7) |
| RIC | 22 (37.3) |
| TBI | 18 (30.5) |
| ATG | 34 (57.6) |
| **GvHD, n (%)** |  |
| *GvHD prophylaxis* |  |
| ATG | 34 (57.6) |
| Calcineurin inhibitors | 58 (98.3) |
| Mycophenolate Mofetil | 32 (54.2) |
| Corticosteroids | 0 (0) |
| Methotrexate | 14 (23.7) |
| Post-transplant cyclophosphamide | 22 (37.3) |
| *History of GvHD between transplantation and inclusion* |  |
| Acute GvHD | 42 (71.2) |
| Grade I/II/III | 28/12/2 |
| Chronic GvHD | 8 (13.6) |
| Grade I/II/III | 3/3/2 |
| *GvHD status at inclusion* |  |
| No history of GvHD | 15 (25.4) |
| Resolved GvHD | 23 (39.0) |
| Active GvHD (acute or chronic) | 21 (35.6) |
| **Immunophenotyping, median [Q1-Q3]** |  |
| Lymphocytes (NV, 1000-2800/µL) | 1210 [790-2220] |
| CD3^+^ T-lymphocytes (NV, 521-1772/µL) | 666 [422.5-1017] |
| CD3^+^ CD4^+^ T-lymphocytes (NV, 336-1126/µL) | 261 [124-345] |
| Naïve CD4^+^ (CD45^+^CCR7^+^) (NV, 121-456/µL) | 16.0 [8.0-33.5] |
| Central memory CD4^+^ (CD45RA^-^CCR7^+^) (NV, 92-341/µL) | 54.5 [21.5-85.8] |
| Effector memory CD4^+^ (CD45RA^-^CCR7^-^) (NV, 59-321/µL) | 129 [87.0-207.8] |
| Differentiated memory CD4^+^ (CD45RA^+^CCR7^-^) (NV, 11-102/µL) | 4.5 [0.0-29.75] |
| CD3^+^ CD8^+^ T-lymphocytes (NV, 125-780/µL) | 366 [223-618] |
| Naïve CD8^+^ (CD45^+^CCR7^+^) (NV, 86-257µL) | 20.0 [10.0-52.0] |
| Central memory CD8^+^ (CD45RA^-^CCR7^+^) (NV, 19-93/µL) | 10.0 [3.3-17.0] |
| Effector memory CD8^+^ (CD45RA^-^CCR7^-^) (NV, 15-162/µL) | 157 [75-281] |
| Differentiated memory CD8^+^ (CD45RA^+^CCR7^-^) (NV, 39-212/µL) | 156 [57.8-309.3] |
| CD4^+^/CD8^+^ ratio (NV, 0.9-6) | 0.57 [0.35-0.98] |
| CD20^+^ B-lymphocytes (NV, 64-593/µL) | 173 [90-379] |
| Immunoglobulin G titers (NV, 7-16 g/L) | 7.5 (6.0-9.9) |
| **Post-transplant immunomodulatory therapy at inclusion, n (%)** |  |
| IS therapy at inclusion# | 19 (32.2) |
| IVIG infusion(s) | 40 (67.8) |
| Time since last IVIG infusion (days), median [IQR] | 128 [89.8-174.5] |
| DLI | 6 (10) |

#Immunosuppressive therapies included ciclosporin (n = 9), tacrolimus (n = 4), corticosteroids (n = 8), and ruxolitinib (n = 3)

*Abbreviations: ATG*, antithymocyte globulin; *CM*, central memory; *CR*, complete remission; *D*, donor; *DM*, differentiated memory; *DLI*, donor lymphocyte infusion; *EM*, effector memory; *GvHD*, graft versus host disease; *IS*, immunosuppressive; *IVIG*, intravenous immunoglobulins; *MAC*, myeloablative conditioning; NK, natural killer; *NV*, normal values; *R*, recipient; *RIC*, reduced-intensity conditioning; *TBI,* total body irradiation.

**Supplementary Table S6. Association between transplant-related factors and Euclidean distance calculated from the HV population centroid to each allo-HSCT recipient projected on the PCA, based on the stimulated TruCulture results obtained from the restricted post-stimulation dataset.**

| **Clinical variable** | **No Euclidean distance median [Q1-Q3] Allo-HSCT** | **Yes Euclidean distance median [Q1-Q3] Allo-HSCT** | ***p.adj (Wilcoxon)*** |
| --- | --- | --- | --- |
| **Ongoing immunosuppressive treatment** | **9.71 [4.91-12.6] n= 40** | **13.77 [11.34-18.43] n= 19** | **0.047** |
| Recipients > 60 years old | 11.8 [8.32-14.48] n= 44 | 8.11 [3.63-14.53] n= 15 | 0.933 |
| Sex - Male | 11.59 [5.55-14.79] n= 25 | 11.34 [7.63-13.77] n= 34 | 0.933 |
| Acute myeloid leukemia | 111.55 [4.93-16.66] n= 28 | 11.36 [7.64-12.82] n= 31 | 0.933 |
| Complete remission* | 17.86 [11.36-18.89]  n= 5 | 11.32 [5.38-13.77] n= 53 | 0.715 |
| Reduced intensity conditioning | 11.59 [5.38-14.79] n= 37 | 11.34 [8.15-14.29] n= 22 | 0.933 |
| Radiotherapy | 11.32 [7.7-14.46] n= 41 | 12.69 [5.81-14.35] n= 18 | 0.933 |
| Anti-thymocyte globulin treatment | 12.22 [3.21-13.77] n= 25 | 11.15 [8.66-14.71] n= 34 | 0.933 |
| Peripheral blood cells | 10.76 [5.34-13.28] n= 12 | 11.36 [7.64-14.63] n= 47 | 0.933 |
| Haploidentical donor | 11.34 [7.76-13.95] n= 48 | 12.52 [4.71-18.37] n= 11 | 0.933 |
| History of acute GvHD  since transplant | 12.1 [7.7-13.77] n= 17 | 11.15 [5.42-14.52] n= 42 | 0.933 |
| History of chronic GvHD  since transplant | 11.59 [5.46-14.67] n= 51 | 11.34 [10.13-12.06] n= 8 | 0.933 |
| T cells CD4+ > 200 cells/µL | 11.79 [6.19-14.29] n= 22 | 10.97 [7.58-14.54] n= 37 | 0.933 |
| T cells CD4+/CD8+ ratio >1 | 12 [7.47-14.95] n= 44 | 9.27 [6.18-12.51] n= 15 | 0.933 |
| CMV reactivation status | 10.48 [5.3-12.86]  n= 46 | 16.26 [11.59-18.89]  n= 13 | 0.948 |

*Data were missing for one patient

*Abbreviations: GvHD*, graft versus host disease

**Supplementary Table S7. Association between transplant-related factors and Euclidean distance calculated from the HV population centroid to each allo-HSCT recipient projected on the PCA, based on the unstimulated PaxGene results.**

| **Clinical variable** | **No Euclidean distance median [Q1-Q3] Allo-HSCT** | **Yes Euclidean distance median [Q1-Q3] Allo-HSCT** | ***p.adj (Wilcoxon)*** |
| --- | --- | --- | --- |
| Ongoing immunosuppressive treatment | 11.97 [8.63-13.83] n= 40 | 10.5 [9.6-15.21] n= 19 | 0.957 |
| Recipients > 60 years old | 11.83 [8.91-13.92] n= 44 | 11.6 [9.19-13.89] n= 15 | 0.957 |
| Sex - Male | 11.6 [9-13.22] 25 | 11.78 [8.87-14.43] n= 34 | 0.957 |
| Acute myeloid leukemia | 11.63 [9.39-14.38] n= 28 | 11.79 [8.83-13.68] n= 31 | 0.957 |
| Complete remission* | 13.22 [11.92-13.83] n= 5 | 10.62 [8.56-13.92] n= 53 | 0.957 |
| Reduced intensity conditioning | 12.15 [10.18-13.92] n= 37 | 9.81 [7.35-13.14] n= 22 | 0.957 |
| Radiotherapy | 11.88 [9.53-13.85] n= 41 | 10.07 [7.32-14.4] n= 18 | 0.957 |
| Anti-thymocyte globulin treatment | 12.06 [9-14.68] n= 25 | 11.26 [8.87-13.75] n= 34 | 0.957 |
| Peripheral blood cells | 12.32 [8.56-14.38] n= 12 | 11.6 [9.09-13.84] n= 47 | 0.957 |
| Haploidentical donor | 11.69 [9.31-13.99] n= 48 | 11.67 [8.28-12.9] n= 11 | 0.957 |
| History of acute GvHD  since transplant | 11.88 [7.96-13.92] n= 17 | 11.63 [9.13-13.75] n= 42 | 0.957 |
| History of chronic GvHD  since transplant | 10.93 [8.6-13.84] n= 51 | 12.57 [11.29-14.3] n= 8 | 0.957 |
| T cells CD4+ > 200 cells/µL | 11.87 [9.71-14.43] n= 22 | 11.67 [7.96-13.5] n= 37 | 0.957 |
| T cells CD4+/CD8+ ratio >1 | 11.83 [9.4-13.87] n= 44 | 10.18 [7.62-14.09] n= 15 | 0.957 |
| CMV reactivation status | 10.78 [7.77-13.85]  n= 46 | 11.95 [10.65-14.12]  n= 13 | 0.957 |

*Data were missing for one patient

*Abbreviations: GvHD*, graft versus host disease

### 2 Supplementary Figure

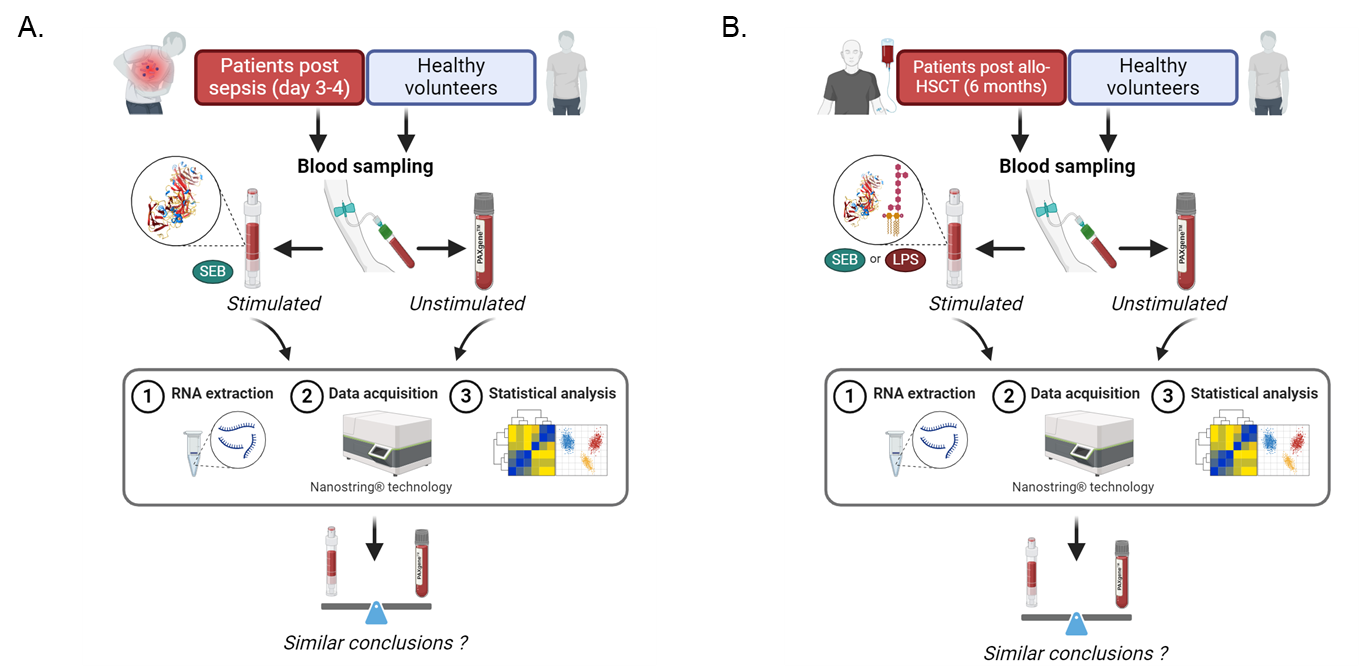

**Supplementary Figure 1.** Graphical representation of the study analysis workflow

Pipeline for the transcriptomic analysis comparing stimulated and unstimulated samples from healthy volunteers and (A) patients with sepsis or (B) allo-HSCT recipients.

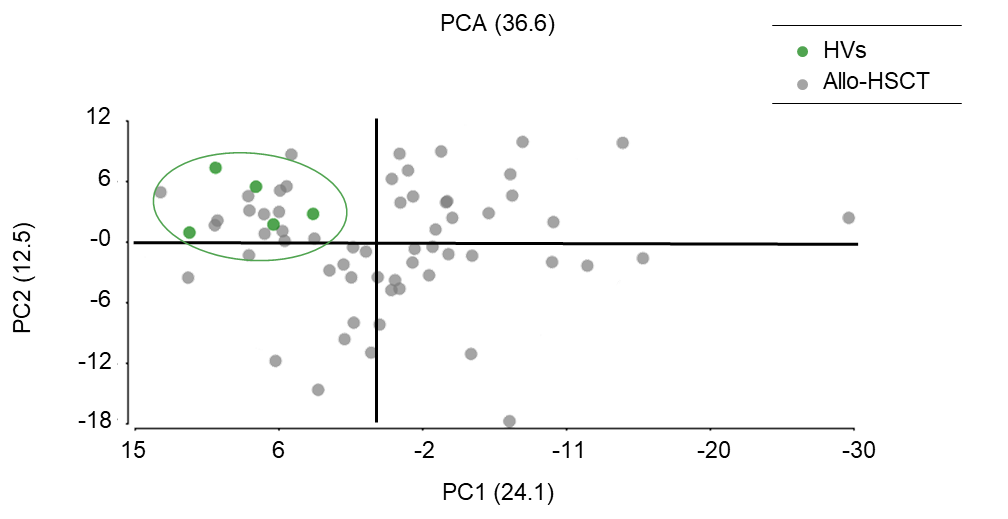
*Abbreviations: LPS,* lipopolysaccharide*; SEB,* Staphylococcal Enterotoxin B

**Supplementary Figure 2.** Individual transcriptomic profile of the allo-HSCT population compared to the HV population from the restricted post-stimulation dataset.

Principal Component Analysis (PCA) of gene expression data derived from 5 HVs (light green) and 59 allo-HSCT recipients (grey) on the restricted dataset post-LPS and –SEB stimulation. Individuals were represented onto the first 2 principal components and each circle represents an individual transcriptomic profile. PCA shows the presence of a specific cluster formed by HV individuals (green ellipse), the HV centroid (green circle) serves as a reference for functional transcriptomic response.

*Abbreviations: HVs*, healthy volunteers, *HSCT*, hematopoietic stem cell transplantation, *LPS*, lipopolysaccharide, *SEB*, staphylococcal enterotoxin B, *PCA*, principal component analysis, *PC*, principal component
